# SLC25A48 is a human mitochondrial choline transporter

**DOI:** 10.1101/2023.12.04.23299390

**Authors:** Suraj Patil, Oleg Borisov, Nora Scherer, Christophe Wirth, Pascal Schlosser, Matthias Wuttke, Kai-Uwe Eckardt, Carola Hunte, Björn Neubauer, Anna Köttgen, Michael Köttgen

## Abstract

Choline has important physiological functions as a precursor for essential cell components and signaling molecules including phospholipids and the neurotransmitter acetylcholine. Choline is a water-soluble charged molecule and therefore requires transport proteins to cross biological membranes. Membrane transport of choline is incompletely understood. Here we show that SLC25A48 is a human mitochondrial choline transporter. Loss-of-function mutations in *SLC25A48* are associated with elevated urine and plasma choline levels resulting from impaired choline transport into mitochondria.

## Main

Membrane transport proteins play a crucial role in the movement of ions and metabolites across biological membranes. Their proper functioning is essential for many physiological processes. Despite significant progress in our understanding of these proteins, a substantial number of them remain uncharacterized, often referred to as orphan membrane transport proteins^1^. Deorphanization efforts are essential to unveil the hidden cellular functions and roles of these proteins in health and disease. Recent large-scale genome-wide association studies of metabolite levels (mGWAS) provide links between common genetic variants in membrane transporter-encoding genes and metabolite levels^2,3^. This generates testable hypotheses to identify the physiological substrates of established and orphan human transport proteins *in vivo*.

Choline is an essential nutrient with important roles in a variety of physiological processes and metabolic pathways. It is an essential component of phospholipids in cell membranes such as phosphatidylcholines, a precursor of the neurotransmitter acetylcholine and of the osmoregulatory betaine, and an important player in lipid metabolism^4^. Recent studies have uncovered a growing body of evidence linking choline to various diseases such as neurological disorders, metabolic syndromes, and liver disease^4^. Several choline transport proteins have been identified and characterized in model systems, including SLC5A7, SLC44A1, SLC44A2, SLC44A4, FLVCR1, and FLVCR2^5–10^. However, it is unclear which transporters affect systemic levels of free choline in humans and if they are linked to human genetic variation.

We previously found through mGWAS that common *SLC25A48* variants associate with altered choline levels in urine (**Fig. 1a**)^2^, and now established that the genetic basis of this association is shared with plasma choline as well as *SLC25A48* transcript levels in brain tissue (**Supplementary** Fig. 1), supporting the hypothesis that the orphan solute carrier SLC25A48 may be a choline transporter. Overexpressed SLC25A48 localizes to mitochondria in human epithelial cells (**Fig. 1b**). We therefore isolated mitochondria of cells over-expressing SLC25A48 and measured mitochondrial uptake of radio-labelled choline (**Fig. 1c**). Mitochondria from cells over-expressing SLC25A48 showed a significant increase of choline uptake compared to mitochondria from control cells (**Fig. 1d**). Mitochondrial uptake of choline was time-and concentration-dependent (**Fig. 1e, f**). These data establish that SLC25A48 is a newly identified, high-affinity mitochondrial choline transporter. SLC25A48 transports choline at nanomolar concentrations (**Fig. 1f**), and thus operates at physiological concentrations of free choline in humans (7-20 µM) ^11^.

**Fig. 1:**
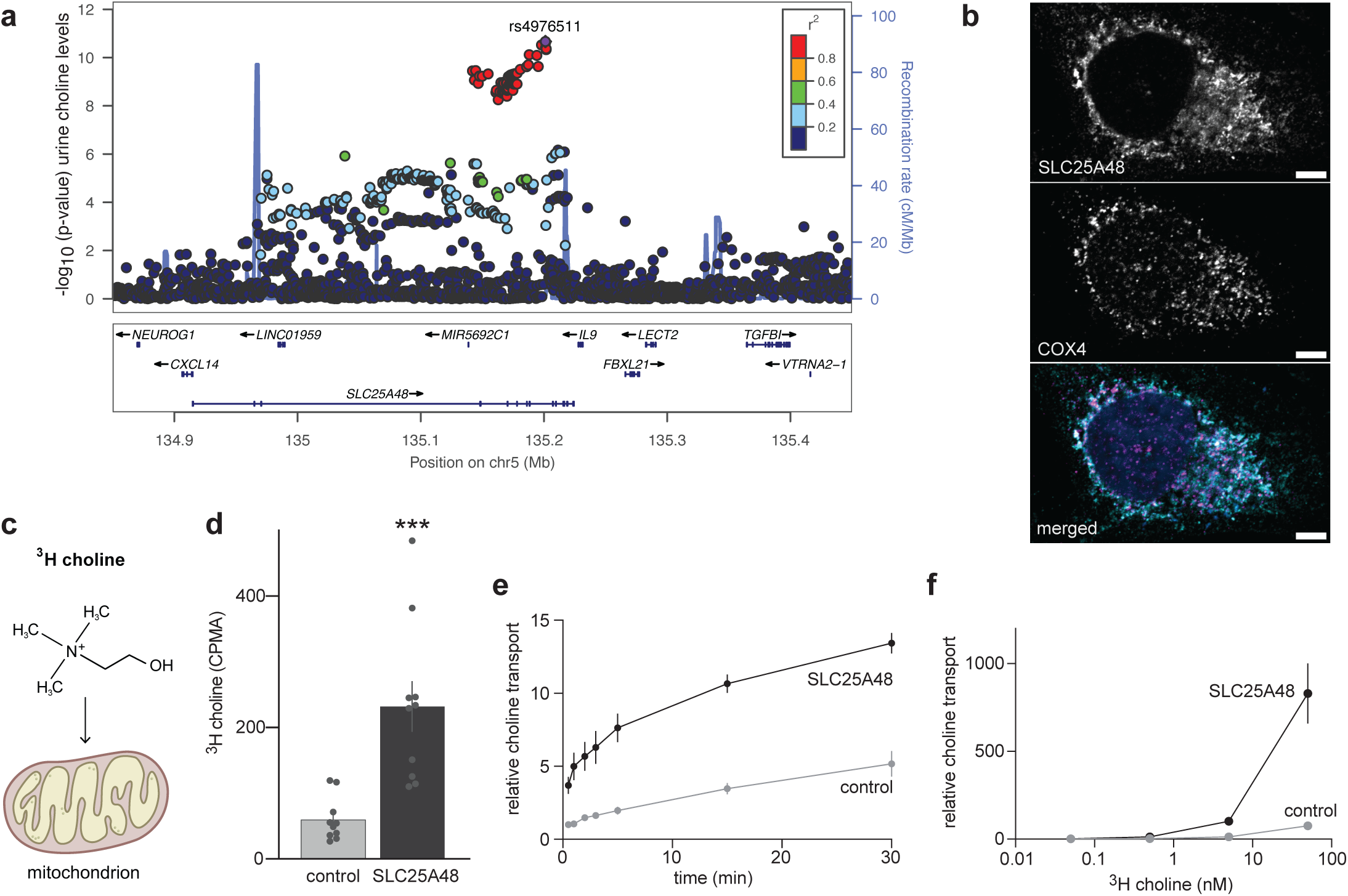
SLC25A48 is a human mitochondrial choline transporter. **(a)** Regional association plot showing association of common genetic variants in the *SLC25A48* locus with choline levels in urine. **(b)** Flag-tagged SLC25A48 localizes to mitochondria as shown by co-localization with the mitochondrial protein COX4. Colors in merged image: SLC25A48 (turquoise), COX4 (violet), and DAPI (blue). scale bar: 5 µM. **(c, d)** Mitochondrial uptake of radio-labelled ^3^H-choline in cells expressing SLC25A48 compared to mock-transfected control cells (n=10). Average counts per minute (CPMA). *** p<0.001. **(e)** Time course of relative choline uptake in mitochondria. **(f)** Concentration-dependence of choline uptake in mitochondria from cells expressing SLC25A48 compared to mock-transfected controls.

To test whether there is a causal link between altered choline levels in humans and impaired mitochondrial choline transport, we investigated the aggregated impact of rare, putatively damaging variants in *SLC25A48* and found significant associations with urine and plasma choline levels (burden test P-values 1.4e-17 and 4.2e-06, respectively; **Supplementary Table 1**). Carriers of driver variants (Methods) showed significantly higher levels of choline compared to non-carriers, which was more pronounced in urine than in plasma (**Fig. 2a**). Identified driver variants mapped into different regions of *SLC25A48* (**Fig. 2b**).

**Fig. 2:**
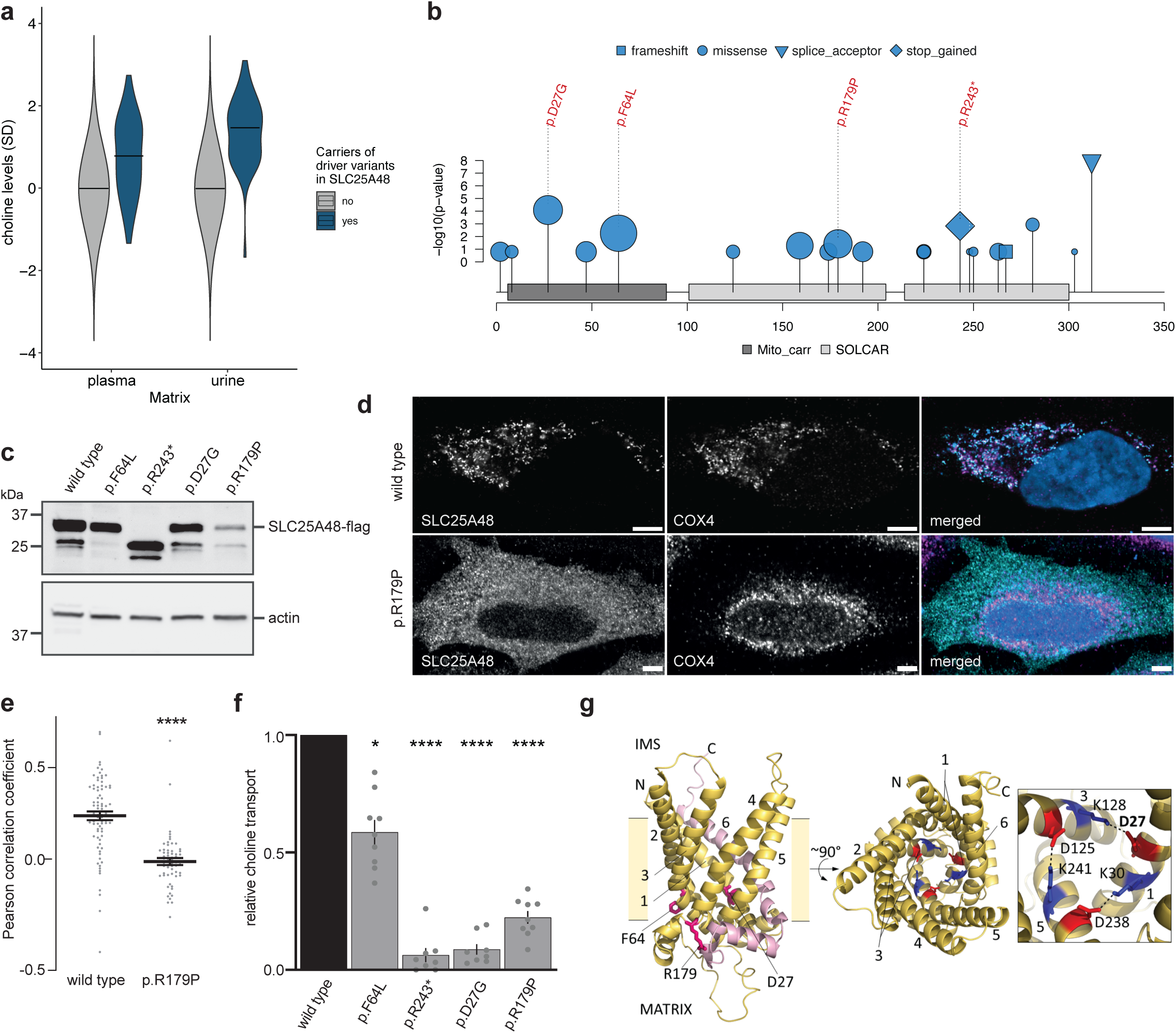
Rare damaging variants in *SLC25A48* impair choline transport. **(a)** Comparison of inverse normal transformed choline levels in urine and plasma among carriers (N=47) and non-carriers (N=5,572 for urine; N=4,666 for plasma) of putative rare damaging driver variants in *SLC25A48* (P-value unpaired t-test (two-tailed): 3.8e-21 for urine and 1.3e-07 for plasma). **(b)** Localization of rare, damaging driver variants with respect to their protein position in SLC25A48 (Q6ZT89 corresponding to transcript ENST00000681962.1, domains based on InterPro, x-axis). Symbol shape corresponds to variant consequence and the size represents the positive effect size of each individual variant on urine choline levels (**Supplementary Table 1**). Individual variant association P-values with urine choline levels are shown on the y-axis. Variants selected for subsequent functional analyses are labeled. **(c)** Western blot analysis of wild type versus mutant flag-tagged SLC25A48. Actin was used as loading control. **(d)** Cellular localization of the SLC25A48 missense mutation p.R179P compared to wild type SLC25A48. Indirect immunofluorescence of SLC25A48-flag and COX4 as mitochondrial marker. Colors in merged image: SLC25A48 (turquoise), COX4 (violet), and DAPI (blue). scale bar: 5 µM. **(e)** Quantification of co-localization of SLC25A48 and COX4 (Pearson correlation coefficient) shows a significant reduction of mitochondrial localization of SLC25A48-R179P compared to wild type (**** p<0.0001; see **Supplementary** Fig. 2 for other mutants). **(f)** Relative mitochondrial uptake of radio-labelled ^3^H-choline in cells expressing wild-type and mutant SLC25A48. Transport of mutant SLC25A48 was normalized to wild-type transport. * p<0.05, **** p<0.0001. **(g)** Position of damaging mutations in model of SLC25A48 (AlphaFold) in intermembrane space (IMS)-facing conformation (helices numbered). Left: Side chains of mutant residues are highlighted in pink. The protein part that would be truncated in p.R243* is colored in pink. Center, right: Residues of the conserved matrix-salt-bridge network (dotted line) including D27 (mutation in p.D27G) are highlighted in red and blue sticks for acidic and basic residues, respectively.

Choline has been implicated in various human conditions ranging from neurological diseases to metabolic traits, but there is limited evidence linking genetic defects in choline transport to human disease. We therefore tested whether choline-related, putative loss-of-function (pLoF) mutations in *SLC25A48* were associated with any of the human traits and diseases ascertained in the UK Biobank that are related to tissues where *SLC25A48* is highly expressed (Methods). Although none of the binary (**Supplementary Table 2**) or quantitative (**Supplementary Table 3**) traits was significantly associated with *SLC25A48* pLoF carrier status after correction for multiple testing, we note several suggestive associations with congenital, musculoskeletal, neurological, and eye diseases or traits.

To investigate whether implicated variants were indeed causally related to altered choline levels, we generated four mutations in the *SLC25A48* cDNA using site-directed mutagenesis and over-expressed these variants in human cell lines to study their effect on protein expression, localization, and function. Western blot analyses showed reduced protein expression of some but not all investigated *SLC25A48* mutations compared to wild-type protein (**Fig. 2c**). Interestingly, some mutations showed mis-localization from mitochondria (p.R179P and p.R243*), with a significant reduction of the mitochondrial co-localization index compared to wild-type SLC25A48 (**Fig. 2d, e**; **Supplementary** Fig. 2), whereas other mutations localized to mitochondria like wild-type transporters (p.D27G and p.R64L; **Supplementary** Fig. 2). These data show that only some of the investigated *SLC25A48* mutations result in reduced protein expression and/or mis-localization.

We subsequently measured choline uptake in mitochondria to investigate which mutations affect SLC25A48 transport function. Despite differences in abundance and localization, all tested SLC25A48 mutations significantly impaired mitochondrial choline uptake (**Fig. 2f**). These data establish a causal link between impaired mitochondrial choline transport via SLC25A48 and altered plasma and urine choline levels in humans. From a mechanistic standpoint, our data suggest that the investigated mutations cause loss-of-function through different pathogenic mechanisms, including reduced expression, mis-localization, and impaired substrate turnover with normal expression level and localization.

To gain a better mechanistic understanding why specific mutations cause impaired transport, we analyzed structural models of SLC25A48 in two conformations (**Fig. 2g, Supplementary** Fig. 3). The SLC25A48 models were in very good agreement with experimental structures of other SLC25 family members, such as the ADP/ATP carrier and mitochondrial uncoupling protein (UCP) (**Supplementary** Fig. 3). Notably, residue D27 is part of a matrix-salt-bridge network of conserved residues, which is a key component of the matrix gate of these transporters and thus essential for transport function. This in line with the strongly impaired choline transport of p.D27G despite its normal localization^12,13^. The other mutations were also localized on the matrix side of the transporter and may affect its folding and stability.

In summary, we show that the physiological function of SLC25A48 in humans is choline import into mitochondria. Loss-of-function mutations in *SLC25A48* impair choline transport into mitochondria, thereby causing higher levels of free choline in urine and plasma. The deorphanization of SLC25A48 defines its molecular function in humans and enables future well-powered studies addressing its role in health and disease.

## Methods

### Molecular biology

pDONR221_SLC25A48 was a gift from the RESOLUTE Consortium (Addgene plasmid # 131995; http://n2t.net/addgene:131995; RRID:Addgene_131995). To incorporate a 5’-Mlu1-and a 3’-Not1-restriction site into the SLC25A48 transcript, PCR was carried out using Pfu Ultra enzyme and the following primers CGC GGG ACG CGT GCC ACC ATG GGC AGC TTC CAG CTG GA and CGC GGG GCG GCC GCC TGG GGA TGT CAC TGC GTG. The resulting fragment was ligated into the pcDNA6.flag vector. To generate plasmids with specific damaging variants, site-directed mutagenesis was performed using the following primers: hSLC25A48.flag _D27G: CTT GTC TTC hSLC25A48.flag_F64L: GCT GGC CAG TGG TAA AGA CAT GCC CTT AAA GAA GCC; GGC TTC TTT AAG GGC ATG TCT TTA CCA CTG GCC AGC; hSLC25A48.flag_R179P: GCT TGC TCC CGG ATA CAG TCC TGC CAG TCC C; GGG ACT GGC AGG ACT GTA TCC GGG AGC AAG C; To obtain the truncated version SLC25A48.flag_R243*, a shorter transcript was derived from the hSLC25A48.flag.pcDNA6 plasmid through PCR with the primers CGC GGG ACG CGT GCC ACC ATG GGC AGC TTC CAG CTG GA and CGC GGG GCG GCC GCC AGA CTT CAC CAC GTC CAT which then was subcloned into pcDNA6.flag vector as described before.

### Cell Culture

HEK293T and HeLa cell lines were obtained from the American Type Culture Collection. Both cell lines were cultured as adherent monolayers in Dulbecco’s Modified Eagle Medium (DMEM, Gibco), supplemented with 10% heat-inactivated fetal bovine serum (FBS, Biochrom) and Penicillin-Streptomycin (Sigma Aldrich P8781) in a humidified incubator with 5% CO2 at a temperature of 36.5°C. Cells were routinely passaged every 3–4 days using 0.05% trypsin-EDTA (Invitrogen) for detachment. For transfection, HEK293T cells were transfected with either 10 µg of plasmid per 55 cm^2^ dish or 100 µg of plasmid per 500 cm^2^ plates using the calcium phosphate transfection method. HeLa cells were transfected using FuGENE HD transfection reagent according to the manufacturer’s instructions (Promega E2311).

### Protein isolation, gel electrophoresis, and Western blot

Cells were harvested 48 hours post transfection, and protein isolation and processing were conducted following established procedures as described previously^14^. Briefly, cell samples were lysed in a cold IP buffer (1% Triton X-100, 20 mM Tris-HCl pH 7.5, 50 mM NaCl, 50 mM NaF, 15 mM Na4P2O7, and 0.1 mM EDTA pH 8) supplemented with 2 mM Na3VO4 and complete protease inhibitor cocktail tablets (Roche 11697498001). The lysates were then subjected to centrifugation at 4°C for 15 minutes at 10,000 g. Supernatants were prepared for further analysis by denaturation for 30 minutes at 42°C with 2x Laemmli buffer.

The denatured protein samples were separated using SDS-PAGE, employing precast Mini-PROTEAN TGX 4-15% gels (BioRad 4561086). The separated proteins were then transferred onto PVDF membranes via a wet blot system for 1 hour at 100 V (BioRad). Membranes were blocked with 5% BSA and subsequently incubated with primary antibodies anti-flag (1:3000; Sigma Aldrich F3165and anti-ß-actin (1:5000; Sigma Aldrich A1978) overnight at 4°C. For detection secondary antibodies (anti-rabbit (GE Healthcare NA934V) and anti-mouse (Dako P0447), were used at a dilution of 1:10,000). Chemiluminescence signals were acquired using the Intas ChemoCam system (Intas Science Imaging) within the dynamic range of the charge-coupled device sensor, ensuring that none of the analyzed bands were saturated. The Western blot in Fig. 2c is representative of three experiments with similar results.

### Immunofluorescence

300,000 Hela cells were seeded in µdish 35mm dishes (ibidi, 81156) and transfected the following day with 2.5 µg of plasmid in 7.8 µl of FuGENE HD transfection reagent (Promega, E2311) and 117.2 µl of water. Two days after transfection, cells were fixed with 3.2 % paraformaldehyde (PFA) for 7 minutes 30 seconds and then permeabilized with a 0.05% Triton-X solution in PBS for 15 minutes. A blocking step was carried out using a mixture of 5% horse serum and 1% BSA in PBS (IF blocking buffer) for 1 hour at RT.

The cells were then incubated overnight with primary antibodies in IF blocking buffer, including anti-flag (Sigma Aldrich F3165, 1:1000) and anti-COX4 (Cell Signaling 4850, 1:200). The following day, the samples were incubated with secondary antibodies for 90 minutes (Hoechst 33342 (Thermo Fisher Scientific, H1399, 1:10000), Alexa Fluor 647 goat anti-rabbit (Invitrogen, A21245, 1:1000), and anti-mouse Alexa Fluor 488 (Molecular Probes, A-11029)). washed, and mounted for microscopy (Dako Glycergel, Agilent). IF-Imaging was performed on a on a LSM980 MP AiryScan 2 equipped with Plan-Apochromat 63x 1.4 oil (Zeiss) within the dynamic range of the detector (pixel size 59 nm x 59 nm). A quantitative colocalization analysis of Flag-tagged SLC25A48 (Alexa Fluor 488) and COX4 (Alexa Fluor 647) was performed with the Colocalization plugin in the Zeiss ZEN blue 3.4 software. 55 to 86 ROIs per group comprising whole cells without nucleus originating from three independent transfections were manually determined. Costes optimal threshold was applied and pixel-based colocalization between two channels was measured to calculate Pearson’s correlation coefficient per ROI.

### Mitochondrial isolation

HEK293T cells were seeded in a 1:12 dilution in 500 cm^2^ plates and transfection was performed the following day. Two days after transfection, cells were harvested to obtain crude mitochondrial pellets, following the protocol described in^15^. The mitochondrial pellets were resuspended in a mitochondrial uptake buffer, which was composed of KCl (120 mM), sucrose (25 mM), HEPES (10 mM), EGTA (1 mM), KH2PO4 (1 mM), MgCl2 (5 mM), glutamate (15 mM), and malate (7.5 mM), adjusted to a pH 7.2 ^6^.

## ^3^Hcholine Transport studies

A total of 25 µg of crude mitochondria was suspended in 50 µl of mitochondrial uptake buffer at room temperature. Subsequently, 50 µl of 2x choline buffer (mitochondrial uptake buffer supplemented with 20 µM choline chloride (Merck C7017-5G) (cold choline)), and 10 nM Choline Chloride, [Methyl-3H]-, 1 mCi (revvity, NET109001MC) (hot choline)), resulting in a final concentration of 10 µM cold choline and 5 nM hot choline. The mixture was incubated at room temperature for 5 minutes to facilitate radiolabeled transport.

Upon completion of the incubation, 1 ml of ice-cold mitochondria washing buffer (mitochondrial uptake buffer containing an additional 20 µM choline chloride) was added, and the samples were promptly placed on ice. Following this, centrifugation was carried out at 10,000 g for 5 minutes at 4°C to remove the radiolabeled choline. The resulting pellet was subjected to an additional wash with 300 µl of mitochondria washing buffer through centrifugation at 10,000 g for 5 minutes at 4°C. The final mitochondrial pellet was resuspended in 4 ml of Ultima Gold scintillation cocktail (revvity 6013329), and the ^3^H-choline content was quantified using liquid scintillation counting.

For time-course experiments, the incubation duration for radiolabeled transport was varied: 30 seconds, 1 minute, 2 minutes, 3 minutes, 5 minutes, 15 minutes, and 30 minutes. For concentration dependence experiments, the choline buffer exclusively contained hot choline (no cold choline) at four different concentrations: 0.05 nM, 0.5 nM, 5 nM, and 50 nM. For time-or concentration-dependent choline uptake experiments, data were normalized to the 30 s timepoint or the 0.5 nM measurement from control samples respectively (Fig. 1e, f). For the quantification of relative choline transport (Fig. 2f), the transport activity of mock-transfected mitochondria was subtracted from the transport activity observed in mitochondria over-expressing wild type and mutant SLC25A48, and normalized to the respective wild type transport activity.

### Statistics

GraphPad Prism® 9.5.1 software was used to graph, analyze and present the obtained data. All results are expressed as mean ± SEM. All experiments were independently performed at least three times and two-tailed Mann-Whitney test was used to calculate p values. A p-value < 0.05 was considered significant.

### Study population

Genetic associations with choline levels were evaluated in the German Chronic Kidney Disease (GCKD) study, an ongoing prospective cohort study of 5,217 participants with chronic kidney disease stages G3; A1-3 or G1-2; A3 at inclusion that has been described in detail before^16,17^. Plasma and urine samples originate from the study’s baseline visit, where biosamples were collected, processed, and shipped frozen to a central biobank for storage at −80 degrees Celsius^18^. The GCKD study was registered in the national registry for clinical studies (DRKS 00003971) and approved by local ethics committees of the participating institutions^16^. All participants provided written informed consent.

### Measurement of choline levels

Choline was measured as part of the non-targeted mass spectrometry-based Metabolon HD4 Global Discovery panel at Metabolon, Inc. Generation and processing of metabolomics data from stored plasma and spot urine samples of the GCKD study has been described previously^2^. Briefly, metabolites were identified by automated comparison of the ion features in the experimental sample to a reference library of chemical standards. Peak quantification was based on the area under the curve, followed by normalization to account for inter-day instrument variation. After quality control, choline levels in urine were normalized for inter-individual dilution using the probabilistic quotient method^19^. Prior to gene-based aggregation testing, choline levels were inverse normal transformed.

### Whole Exome Sequencing (WES)

The *SLC25A48* gene was investigated using WES data of GCKD participants, which has been described previously^20^. Briefly, extracted genomic DNA underwent paired-end 100-bp WES at Human Longevity Inc, using the IDT xGen v1 capture kit on the Illumina NovaSeq 6000 platform. The average coverage of the consensus coding sequence (CCDS) release 22 was 141-fold read depth. Exomes were processed in a custom-built cloud compute platform using the Illumina DRAGEN Bio-IT Platform Germline Pipeline v3.0.7 at Astra Zeneca’s Centre for Genomics Research, including alignment to the GRCh38 reference genome and variant calling^21^.

Sample quality control comprised removal of duplicates, samples with withdrawn consent, mismatch of genetic and reported sex, gonosomal aneuploidies, estimated VerifyBamID contamination level >4%, <94.5% of CCDS bases covered with ≥10-fold coverage, highly related samples (kinship >0.884 by KING --kinship v2.2.3)^21^, and missing sample call rate >0.03. Only samples with available high-quality DNA microarray genotype data and without outlying values (>8 SD) along any of the first 10 genetic principal components from a PCA were retained, for a final sample size of 4,779 samples.

Variant quality control comprised exclusion for coverage of <10x, genotype quality score (GQ) <30, mapping quality score (MQ) <40, quality score (QUAL) <30, read position rank sum score (RPRS) <-2, mapping quality rank sum score (MQRS) <-8, heterozygous variants with a one-sided binomial exact test P-value for Hardy-Weinberg equilibrium of <1e-6 or genotype called based on an alternative allele read ratio <0.2 or >0.8, single nucleotide variants with a Fisher’s strand bias score (FS) >60 and insertions and deletions (indel) with a FS >200, variants that did not pass the DRAGEN calling algorithm filters, and variants with a missing call rate >10% among all remaining samples.

### Variant annotation and rare variant aggregation (burden) testing

Called variants were annotated using Variant Effect Predictor (VEP)^22^ version 109 with standard settings. Predicted deleteriousness of variants was added via REVEL^23^ (version 2020-5) and CADD^24^ (version 3.0) VEP plugins and via dbNSFP version 4.1a for additional prediction scores^25^. The LoFtee VEP plugin^26^ (version 2020-8) was used to downgrade loss-of-function variants. Only variants in transcript ENST00000681962.1, annotated as the MANE Select and Ensembl Canonical transcript for *SLC25A48* by VEP, were considered.

The burden test is appropriate when all variants are assumed to affect the trait in the same direction, as is the case for pLoF variants^27^. Qualifying variants in *SLC25A48* for aggregation in burden tests were selected based on variant frequency and VEP annotations. Two complementary masks of selected variants were evaluated: while both masks only assessed rare variants with MAF of <1% in the MANE Select transcript ENST00000681962.1, they differed in the selection of predicted variant effect. The “LoF_mis” mask contained all variants that were predicted to be either high-confidence loss-of-function variants or missense variants with a MetaSVM score >0 or in-frame non-synonymous variants with a fathmm-XF-coding score >0.5, whereas the “HI_mis” mask contained all variants that were predicted either to have a high-impact consequence defined by VEP (transcript ablation, splice acceptor variant, splice donor variant, stop gained, frameshift variant, stop lost, start lost, and transcript amplification) or to be missense variants with either a REVEL score >0.5, a CADD PHRED score >20, or a M-CAP score >0.025. Burden tests were carried out as implemented in the seqMeta R-package version 1.6.7^28^, adjusting for age, sex, ln(eGFR), the first three genetic principal components as well as serum albumin for plasma choline and ln(UACR) for urine choline, respectively. Genotypes were coded as number of copies of the rare allele (0, 1, 2). Statistical significance was defined as p<0.05. Single-variant association tests between each selected variant and choline levels were performed under additive modeling, adjusting for the same covariates. To prioritize selected rare putatively damaging variants according to their contribution to the gene signal, we used a previously described forward selection procedure^29^: variants *v* were ranked by the magnitude of the difference *Δv*=*Pv* - *P*, where *Pv* corresponds to the p-value of the burden test aggregating all variants except for the variant *v*, and *P* to the total P-value of the burden test including all selected variants. Variants with the greatest *Δv* providing the lowest p-value when aggregated are referred to as “driver variants”. Lower association p-values were observed with the “HI_mis” mask as compared to the ”LoF_mis” mask, suggesting that the former better captures the genetic architecture of *SLC25A48* with respect to choline levels. Therefore, results are shown for the “HI_mis” mask.

## Associations with human traits and diseases

### Colocalization analysis

We performed colocalization analysis to investigate whether genetic variants associated with choline levels were also associated with gene expression levels (i.e., if these variants act as expression quantitative trait loci, eQTLs). Summary statistics for eQTLs were obtained from the Genotype-Tissue Expression Project (GTEx, V8)^30^. We focused on *SLC25A48* expression in tissues where *SLC25A48* was tissue-enhanced according to the Human Protein Atlas^31^: brain, kidney, and skeletal muscle. For colocalization we used the “coloc” R package and applied the enumeration approach implemented in the “coloc.abf” function^32^. This approach tests whether two traits share the same causal variant in a region by estimating posterior probabilities for five hypotheses: both traits have no causal variant in a region (hypothesis H0), either the first or the second trait has a causal variant (H1 and H2, respectively), both traits have causal variants but they are distinct (H3), and both traits share a common causal variant (H4). Posterior probabilities are assigned to each hypothesis and their sum is equal to 1. Two traits show evidence for genetic colocalization when hypothesis H4 obtains the highest probability (0.5 or higher).

### Rare variant collapsing analysis

We performed phenome-wide association analysis of rare variants in *SLC25A48* with binary traits (clinical diagnoses based on ICD-10 codes) and quantitative traits in the UK Biobank, which comprises up to 500,000 individuals with WES and comprehensive phenotypic data^33^ (UK Biobank application ID 64806). We excluded strongly related individuals (for whom ten or more third-degree relatives were identified) and individuals not included in the kinship inference process. Participants of all ancestries were included into analysis. To adjust for population stratification, we included the first 10 principal components based on genotype array data (UKB Data-Field 22009) when performing all downstream regression analyses for both binary and quantitative traits. In addition to principal components, we included sex, age at recruitment, and interaction between sex and age (sex*age) as covariates.

After individual-level filtering, a total of 468,292 individuals were available for association analyses. Both masks were tested: “LoF_mis” (94 variants) and “HI_mis” (191 variants)^20^. All variants in each mask passed the “90pct10dp” quality filter, defined as a read depth of at least 10 in at least 90% of all genotypes for a given variant independent of variant allele zygosity. Carriers and non-carriers of variants in each mask were compared using the collapsing approach, in which carriers are defined as individuals with either heterozygous or alternative homozygous genotype for at least one of the variants in the mask. The remaining individuals were defined as non-carriers. All analyses were performed on the UK Biobank Research Analysis Platform.

### Analysis of binary traits and diseases

Traits and diseases were defined based on ICD-10 codes (International Classification of Diseases, 10th Revision). We queried all distinct diagnosis codes from the UK Biobank database for each participant across all hospital inpatient records in either the primary or secondary position (UK Biobank Data-Field 41270). Next, we grouped ICD-10 codes using phecode system^34^. The phecode system maps related ICD-10 codes to a larger group of codes known as “phecode”. In total, there are approximately 1,500 phecodes. Each phecode is assigned to a clinically meaningful category (e.g., neurological diseases, musculoskeletal diseases) to facilitate interpretation. We analyzed 658 phecodes belonging to the following categories: circulatory system, congenital anomalies, digestive, endocrine/metabolic, genitourinary, mental disorders, musculoskeletal, neurological, pregnancy complications, and retinal traits from sense organs. For each phecode, we defined case-control status of participants based on the presence or absence of the respective phecode. Since we focused on rare variants, we applied Firth’s logistic regression for association testing (mean bias-reducing adjusted scores approach) as implemented in the “brglm2” R package^35^. Firth’s regression provides bias-reduction in case of rare events and also allows for inclusion of covariates. We report 83 phecodes from both masks (73 unique phecodes) with nominally significant associations (P-value <0.05) in **Supplementary Table 2**.

### Analysis of quantitative traits

Among quantitative traits available in the UK Biobank, we selected a subset of 1,143 physical measures, blood assays, and imaging traits that belonged to the following categories: Chapter V Mental and behavioural disorders, Chapter VI Diseases of the nervous system, Chapter VII Diseases of the eye and adnexa, Chapter IX Diseases of the circulatory system, Chapter XI Diseases of the digestive system, Chapter XIII Diseases of the musculoskeletal system and connective tissue, Chapter XIV Diseases of the genitourinary system^21^, as well as all available blood biochemistry measurements from all chapters. Values of these traits were inverse-normal transformed prior to analyses. We applied linear regression for association testing between quantitative traits and collapsed variants adjusting for covariates as mentioned above. We report 106 quantitative traits from both masks (103 unique traits) with nominally significant associations (P-value <0.05) in **Supplementary Table 3**.

### Structural model generation and analysis

The SLC25A48 AlphaFold^36^ model was retrieved from the AlphaFold Protein Structure Database^37^ hosted by EMBL-EBI (https://alphafold.ebi.ac.uk, accession code Q6ZT89). To identify potential homologous proteins with experimental structures available, a sequence-based BLAST^38^ search of the Protein Data Bank as well as a structural homology based search using the DALI Server^39^ were performed. Structures of other SLC25 family members with accession codes 1ock (ADP/ATP carrier from bovine^40^), 2lck (UCP2 from mouse^41^), 4c9g (ADP/ATP carrier 2 from yeast^42^), 4c9j (ADP/ATP carrier 3 from yeast^42^), 6gci (ADP/ATP carrier from *Thermothelomyces* thermophilus^43^), 8g8w (UCP1 from human, GTP bound^12^) and 8hbv (UCP1 from human, nucleotide-free state^44^) were obtained from the Protein Data Bank. Structures and model were superimposed using Coot^45^. As the SLC25A48 AlphaFold model is in the intermembrane space open conformation, Swiss-Model^46^ was used to model the matrix open conformation using the structure of ADP/ATP carrier from *T. thermophilus*^43^ in matrix open conformation as template. Figures were prepared using the PyMOL Molecular Graphics System, Schrödinger, LLC.

## Data Availability

All data produced in the present work are contained in the manuscript

## Acknowledgements

The cDNA for SLC25A48 was kindly provided by the RESOLUTE consortium (https://re-solute.eu). The authors acknowledge Simone Diederichsen and Andreas Ungi for expert technical assistance and graphic support. We would like to thank the Lighthouse Core Facility for assistance with microscopy and cell sorting. The work of N.S., C.H., B.N., A.K. and M.K. was funded by German Research Foundation (DFG) project ID 431984000 (SFB 1453). N.S. was supported by DFG KO 3598/4-2 (to A.K.). M.K. was supported by German Research Foundation (DFG) project ID 239283807 (TRR 152). Germany’s Excellence Strategy (CIBSS, EXC-2189, project ID 390939984) supported the work of C.H., A.K., and M.K.. S.P. was funded by H2020 MSCA-ITN-2019 ID:860977 (TrainCKDis). The work of P.S. was supported by DFG Project-ID 523737608 (SCHL 2292/2-1). Genotyping and urine metabolomics in the GCKD study were supported by Bayer Pharma. Plasma metabolomics has received funding from the Innovative Medicines Initiative 2 Joint Undertaking (JU) under grant agreement no. 115974. The JU receives support from the European Union’s Horizon 2020 research and innovation program and the EFPIA and the JDRF. Any dissemination of results reflects only the authors’ view; the JU is not responsible for any use that may be made of the information it contains. The GCKD study was and is supported by the BMBF (FKZ 01ER 0804, 01ER 0818, 01ER 0819, 01ER 0820 and 01ER 0821) and the KfH Foundation for Preventive Medicine. Unregistered grants to support the study were provided by corporate sponsors (listed at https://gckd.org). We are grateful for the willingness of the patients to participate in the GCKD study. The effort of the study personnel of the various regional centers is highly appreciated. We thank the large number of nephrologists who provide routine care for the patients and collaborate with the GCKD study.

## References

1. Meixner, E. et al. A substrate-based ontology for human solute carriers. Mol Syst Biol 16, e9652 (2020).

2. Schlosser, P. et al. Genetic studies of paired metabolomes reveal enzymatic and transport processes at the interface of plasma and urine. Nat Genet 55, 995–1008 (2023).

3. Suhre, K. et al. Human metabolic individuality in biomedical and pharmaceutical research. Nature 477, 54–60 (2011).

4. Ueland, P.M. Choline and betaine in health and disease. J Inherit Metab Dis 34, 3–15 (2011).

5. Apparsundaram, S., Ferguson, S.M., George, A.L., Jr. & Blakely, R.D. Molecular cloning of a human, hemicholinium-3-sensitive choline transporter. Biochem Biophys Res Commun 276, 862–7 (2000).

6. Bennett, J.A. et al. The choline transporter Slc44a2 controls platelet activation and thrombosis by regulating mitochondrial function. Nat Commun 11, 3479 (2020).

7. Cater, R.J., et al. Structural and molecular basis of choline uptake into the brain by FLVCR2. bioRxiv (2023).

8. Kenny, T.C. et al. Integrative genetic analysis identifies FLVCR1 as a plasma-membrane choline transporter in mammals. Cell Metab 35, 1057–1071 e12 (2023).

9. Michel, V. & Bakovic, M. The solute carrier 44A1 is a mitochondrial protein and mediates choline transport. FASEB J 23, 2749–58 (2009).

10. Traiffort, E., O’Regan, S. & Ruat, M. The choline transporter-like family SLC44: properties and roles in human diseases. Mol Aspects Med 34, 646–54 (2013).

11. Garcia, E. et al. Quantification of choline in serum and plasma using a clinical nuclear magnetic resonance analyzer. Clin Chim Acta 524, 106–112 (2022).

12. Jones, S.A. et al. Structural basis of purine nucleotide inhibition of human uncoupling protein 1. Sci Adv 9, eadh4251 (2023).

13. Ruprecht, J.J. & Kunji, E.R.S. The SLC25 Mitochondrial Carrier Family: Structure and Mechanism. Trends Biochem Sci 45, 244–258 (2020).

14. Hofherr, A. et al. The mitochondrial transporter SLC25A25 links ciliary TRPP2 signaling and cellular metabolism. PLoS Biol 16, e2005651 (2018).

15. Wieckowski, M.R., Giorgi, C., Lebiedzinska, M., Duszynski, J. & Pinton, P. Isolation of mitochondria-associated membranes and mitochondria from animal tissues and cells. Nat Protoc 4, 1582–90 (2009).

16. Eckardt, K.U. et al. The German Chronic Kidney Disease (GCKD) study: design and methods. Nephrol Dial Transplant 27, 1454–60 (2012).

17. Titze, S. et al. Disease burden and risk profile in referred patients with moderate chronic kidney disease: composition of the German Chronic Kidney Disease (GCKD) cohort. Nephrol Dial Transplant 30, 441–51 (2015).

18. Prokosch, H.U. et al. Designing and implementing a biobanking IT framework for multiple research scenarios. Stud Health Technol Inform 180, 559–63 (2012).

19. Dieterle, F., Ross, A., Schlotterbeck, G. & Senn, H. Probabilistic quotient normalization as robust method to account for dilution of complex biological mixtures. Application in 1H NMR metabonomics. *Anal Chem* **78**, 4281-90 (2006).

20. Pfau, A. et al. SLC26A1 is a major determinant of sulfate homeostasis in humans. J Clin Invest 133(2023).

21. Wang, Q. et al. Rare variant contribution to human disease in 281,104 UK Biobank exomes. Nature 597, 527–532 (2021).

22. McLaren, W. et al. The Ensembl Variant Effect Predictor. Genome Biol 17, 122 (2016).

23. Ioannidis, N.M. et al. REVEL: An Ensemble Method for Predicting the Pathogenicity of Rare Missense Variants. Am J Hum Genet 99, 877–885 (2016).

24. Rentzsch, P., Witten, D., Cooper, G.M., Shendure, J. & Kircher, M. CADD: predicting the deleteriousness of variants throughout the human genome. Nucleic Acids Res 47, D886–D894 (2019).

25. Liu, X., Li, C., Mou, C., Dong, Y. & Tu, Y. dbNSFP v4: a comprehensive database of transcript-specific functional predictions and annotations for human nonsynonymous and splice-site SNVs. Genome Med 12, 103 (2020).

26. Karczewski, K.J. et al. The mutational constraint spectrum quantified from variation in 141,456 humans. Nature 581, 434–443 (2020).

27. Lee, S., Abecasis, G.R., Boehnke, M. & Lin, X. Rare-variant association analysis: study designs and statistical tests. Am J Hum Genet 95, 5–23 (2014).

28. Voorman, A., Brody, J., Chen, H., Lumley, T. & Davis, B.. seqMeta: Meta-Analysis of Region-Based Tests of Rare DNA Variants. (2017).

29. Bomba, L. et al. Whole-exome sequencing identifies rare genetic variants associated with human plasma metabolites. Am J Hum Genet 109, 1038–1054 (2022).

30. Consortium, G.T. The GTEx Consortium atlas of genetic regulatory effects across human tissues. Science 369, 1318–1330 (2020).

31. Uhlen, M. et al. Proteomics. Tissue-based map of the human proteome. Science 347, 1260419 (2015).

32. Giambartolomei, C. et al. Bayesian test for colocalisation between pairs of genetic association studies using summary statistics. PLoS Genet 10, e1004383 (2014).

33. Bycroft, C. et al. The UK Biobank resource with deep phenotyping and genomic data. Nature 562, 203–209 (2018).

34. Wu, P. et al. Mapping ICD-10 and ICD-10-CM Codes to Phecodes: Workflow Development and Initial Evaluation. JMIR Med Inform 7, e14325 (2019).

35. Kosmidis, I., Kenne Pagui, E.C. & Sartori, N. Mean and median bias reduction in generalized linear models.. Stat Comput 43–59 (2020).

36. Jumper, J. et al. Highly accurate protein structure prediction with AlphaFold. Nature 596, 583–589 (2021).

37. Varadi, M. et al. AlphaFold Protein Structure Database: massively expanding the structural coverage of protein-sequence space with high-accuracy models. Nucleic Acids Res 50, D439–D444 (2022).

38. Altschul, S.F., Gish, W., Miller, W., Myers, E.W. & Lipman, D.J. Basic local alignment search tool. J Mol Biol 215, 403–10 (1990).

39. Holm, L., Laiho, A., Toronen, P. & Salgado, M. DALI shines a light on remote homologs: One hundred discoveries. Protein Sci 32, e4519 (2023).

40. Pebay-Peyroula, E. et al. Structure of mitochondrial ADP/ATP carrier in complex with carboxyatractyloside. Nature 426, 39–44 (2003).

41. Berardi, M.J., Shih, W.M., Harrison, S.C. & Chou, J.J. Mitochondrial uncoupling protein 2 structure determined by NMR molecular fragment searching. Nature 476, 109–13 (2011).

42. Ruprecht, J.J. et al. Structures of yeast mitochondrial ADP/ATP carriers support a domain-based alternating-access transport mechanism. Proc Natl Acad Sci U S A 111, E426–34 (2014).

43. Ruprecht, J.J. et al. The Molecular Mechanism of Transport by the Mitochondrial ADP/ATP Carrier. Cell 176, 435–447 e15 (2019).

44. Kang, Y. & Chen, L. Structural basis for the binding of DNP and purine nucleotides onto UCP1. Nature 620, 226–231 (2023).

45. Emsley, P., Lohkamp, B., Scott, W.G. & Cowtan, K. Features and development of Coot. *Acta Crystallogr D Biol Crystallogr* 66, 486–501 (2010).

46. Waterhouse, A. et al. SWISS-MODEL: homology modelling of protein structures and complexes. Nucleic Acids Res 46, W296–W303 (2018).

