## Supplementary material for "SLC25A48 is a human mitochondrial choline transporter": suppliment figures and tables

**Affiliations:**
[1] Department of Medicine IV - Nephrology and Primary Care, Faculty of Medicine and Medical Center, University of Freiburg, Freiburg, Germany
[2] Institute of Genetic Epidemiology, Faculty of Medicine and Medical Center - University of Freiburg, Freiburg, Germany
[3] Institute of Biochemistry and Molecular Biology, ZBMZ, Faculty of Medicine, University of Freiburg, Freiburg, Germany.
[4] Department of Nephrology and Medical Intensive Care, Charité - Universitätsmedizin Berlin, Germany
[5] Department of Nephrology and Hypertension, University Hospital Erlangen, Friedrich-Alexander-Universität Erlangen-Nürnberg, Erlangen, Germany
[6] Spemann Graduate School of Biology and Medicine (SGBM), University of Freiburg, Freiburg, Germany
[7] CIBSS - Centre for Integrative Biological Signalling Studies, Albert-Ludwigs-University Freiburg, Freiburg, Germany
[8] BIOSS-Centre for Biological Signalling Studies, University of Freiburg, Freiburg, Germany.
[9] Faculty of Biology, University of Freiburg, Germany

### Supplementary Materials

### Supplementary Figures

**Supplementary Figure 1: Urine choline levels share an underlying genetic signal with plasma choline levels and *SLC25A48* transcript levels in brain.**

**(a**) Regional association plot showing association of common genetic variants in the *SLC25A48* locus with choline levels in plasma. **(b)** Regional association plot showing association of common genetic variants in the *SLC25A48* locus with SLC25A48 transcript levels in the nucleus accumbens (basal ganglia) brain tissue from GTEx V8 (Methods). In both panels, the SNP with the lowest association p-value is labeled. Colocalization testing (Methods) confirmed the same genetic signal as underlying the association of urine choline levels (Figure 1a) and plasma choline levels (posterior probability of a shared signal (H4) = 0.97) as well as *SLC25A48* transcript levels (posterior probability of a shared signal (H4) = 0.93). In both panels, the genetic variant with the lowest association p-value with *SLC25A48* transcript levels in brain (rs12522259) is labeled.

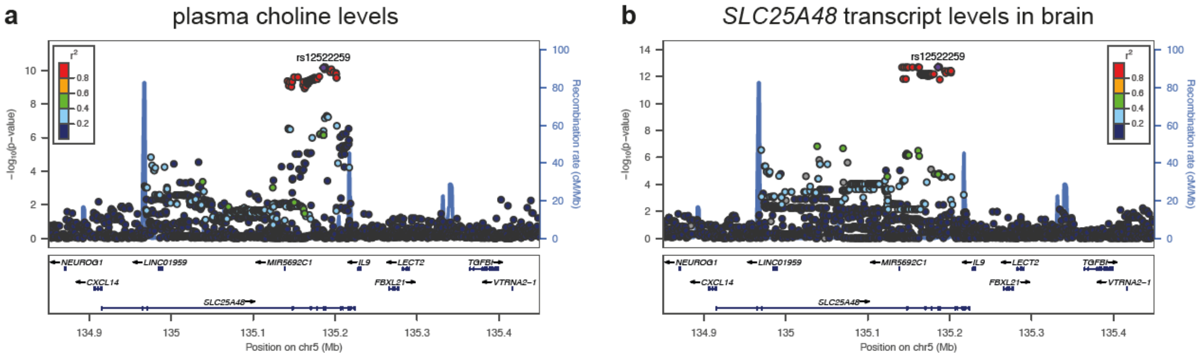

**Supplementary Figure 2: Localization of rare damaging SLC25A48 variants. (a)** Cellular localization of the SLC25A48 missense mutation p.F64L, p.R243*, and p.D27G compared to wild type SLC25A48. Indirect immunofluorescence of SLC25A48-flag and COX4 as mitochondrial marker. Colours in merged image: SLC25A48 (turquoise), COX4 (violet), and DAPI (blue). scale bar: 5 µM. **(b)** Quantification of co-localization of SLC25A48 and COX4 (Pearson correlation coefficient) shows a significant reduction of mitochondrial localization of SLC25A48-R243* compared to wild type (**** p<0.0001). SLC25A48-F64L and SLC25A48-D27G show similar mitochondrial localization like wild type.

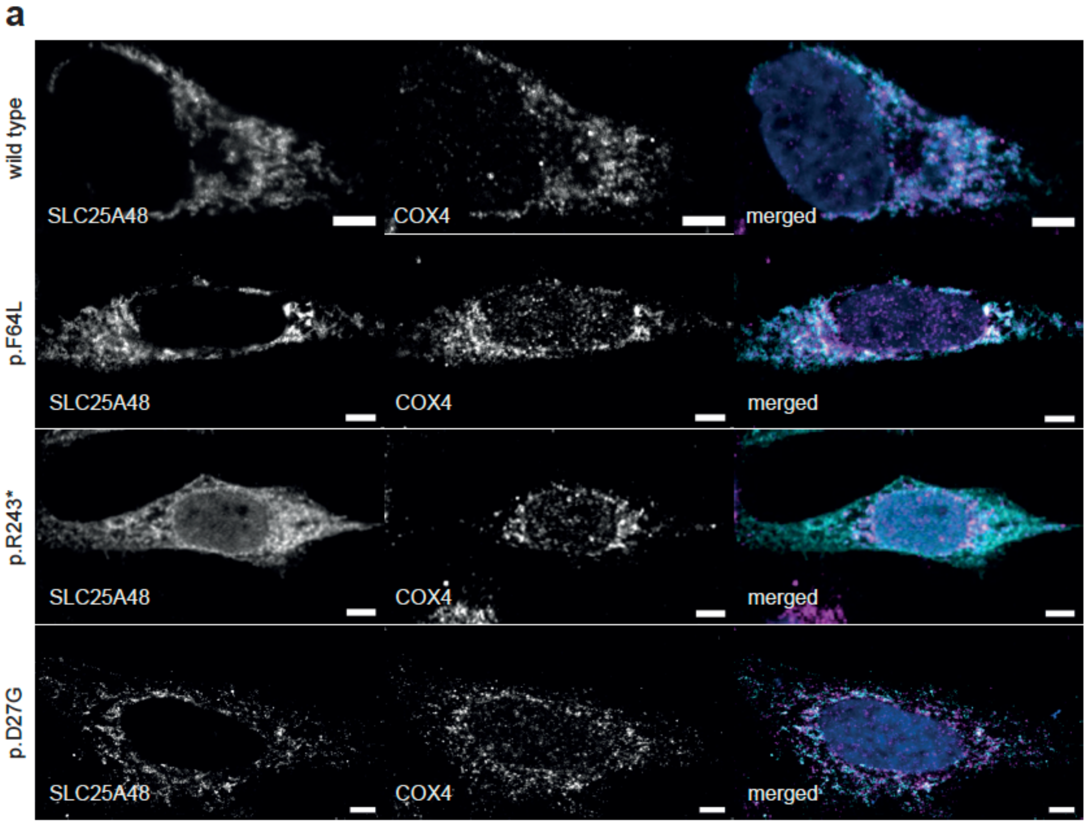

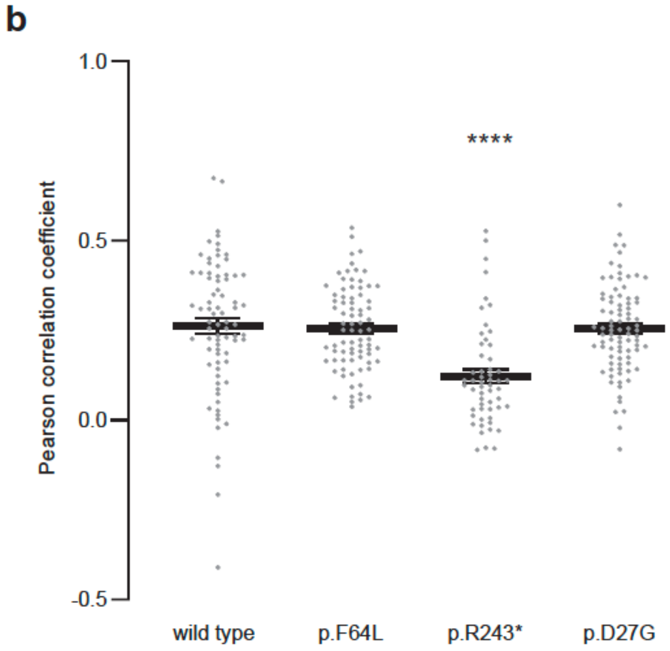

**Supplementary Fig. 3: SLC25A48 models and location of residues affected by specific mutations.**

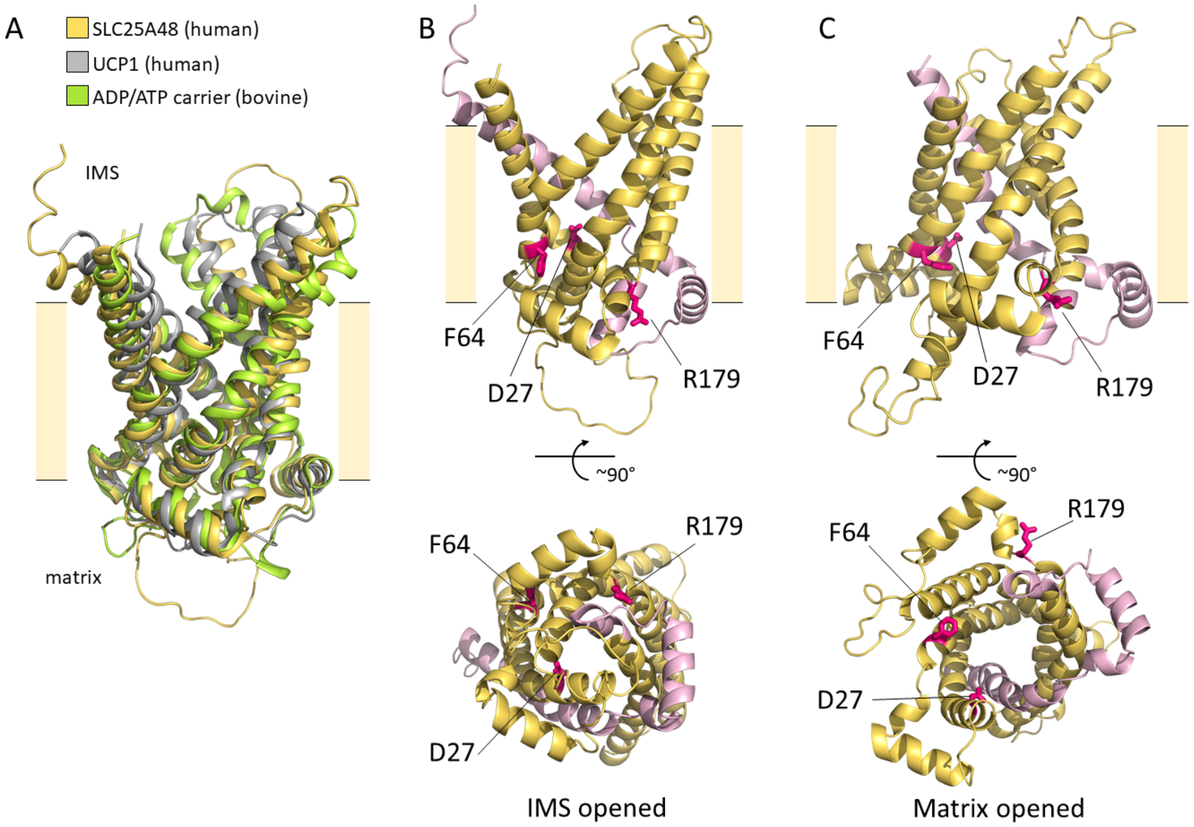

**Supplementary Fig. 3: SLC25A48 models and location of residues affected by specific mutations. (a)** Superposition of SLC25A48 model (AlphaFold Protein Structure database ID Q6ZT89) represented as a yellow cartoon, with two experimental structures of other SLC25 family members. Human UCP1 (PDB code: 8hbv)[1] is shown in grey and bovine ADP/ATP carrier (PDB code: 1okc)[2] is colored in green. The core root-mean-square deviations calculated using the SSM superimposition tool in Coot was 2.7528 Å for 258 aligned residues between SLC25A48 and human UCP1 and 2.4802 Å for 254 aligned residues between SLC25A48 and the bovine ADP/ATP carrier. **(b)** AlphaFold model of SLC25A48 in the intermembrane-space open conformation represented as a cartoon, with the residues affected by the mutations of interest highlighted as in **Fig. 2g**. On the top, the transporter is seen from the membrane plane, with a point of view rotated roughly 90° along the central funnel compared to **Fig. 2g**. On the lower panel, the model is seen from the matrix side with its closed matrix gate. Note that the transporter, as others members of the SLC25 family is formed by three repeats comprising each two transmembrane helices (TMH) connected by a short helix on the matrix side. **(c)** Model of SLC25A48 in the matrix-open conformation, as generated by Swiss-Model using the structure of the ADP/ATP carrier from *T. thermophilus*[3] in matrix-open conformation, as template. Mutations and orientations are as in **(b)**. In this conformation, the transporter is open toward the matrix with a significant enlargement of the transporter diameter on this side. Besides the D27G mutation described in the main text, the truncation at residue 243 has obvious consequences for the fold and function of SLC25A48, as the transporter is missing a connecting helix on the matrix side as well as an entire TMH. The R179P mutation affects a region with high sequence conservation levels. In the models, the residue points toward an adjacent loop and the lipid bilayer where the positively charged side chain of R179 might be required for interactions with phospholipids. It is likely that a mutation to proline at R179 affects the stability and the SLC25A48 transport cycle, in line with the observed mislocalization and reduced transport activity. Interestingly, residue F64 is located in TMH2 and points, in the SLC25A48 model with the matrix-open conformation, toward K128 and Q132, two key residues involved of the opened matrix gate[4]. Loss of the aromatic ring in the F64L variant could weaken the interaction with those neighboring residues in that conformation (in particular in case of a possible cation-π interaction with K128). Overall, the models support the experimental data and suggest that the mutations of interest are likely to affect SLC25A48 stability, folding and transport mechanism.

**References:**

1. Kang, Y. & Chen, L. Structural basis for the binding of DNP and purine nucleotides onto UCP1. *Nature* **620**, 226-231 (2023).
2. Pebay-Peyroula, E. *et al.* Structure of mitochondrial ADP/ATP carrier in complex with carboxyatractyloside. *Nature* **426**, 39-44 (2003).
3. Ruprecht, J.J. *et al.* The Molecular Mechanism of Transport by the Mitochondrial ADP/ATP Carrier. *Cell* **176**, 435-447 e15 (2019).
4. Ruprecht, J.J. & Kunji, E.R.S. The SLC25 Mitochondrial Carrier Family: Structure and Mechanism. *Trends Biochem Sci* **45**, 244-258 (2020).

### Supplementary Tables

**Supplementary Table 1: Rare, putative damaging variants in *SLC25A48* identified in the GCKD study (HI_mis mask) and their associations with urine choline levels.**

**Supplementary Table 2: Results from collapsing tests of the effect of putative loss-of-function variants in *SLC25A48* on binary human traits and diseases in the UK Biobank.**

**Supplementary Table 3: Results from collapsing tests of the effect of putative loss-of-function variants in *SLC25A48* on quantitative human traits in the UK Biobank.**

All Supplementary Tables are provided in a separate spreadsheet, "Patil_SLC25A48_SupplTables_20231111.xlsx".

**Supplementary Table 1: Rare, putative damaging variants in *SLC25A48* identified in the**

| Gene ID | Gene | Chr | Variant | Existing variation | Position (b38) |
|---|---|---|---|---|---|
| ENSG00000145832 | *SLC25A48* | 5 | 5:135852592:C:A | COSV50849439 | 135852592 |
| ENSG00000145832 | *SLC25A48* | 5 | 5:135874068:C:T | rs992362233 | 135874068 |
| ENSG00000145832 | *SLC25A48* | 5 | 5:135842449:A:G | rs1168702512 | 135842449 |
| ENSG00000145832 | *SLC25A48* | 5 | 5:135871575:G:C | rs371126144 | 135871575 |
| ENSG00000145832 | *SLC25A48* | 5 | 5:135871515:A:T | rs1052163123 | 135871515 |
| ENSG00000145832 | *SLC25A48* | 5 | 5:135850474:G:T | rs373533677 | 135850474 |
| ENSG00000145832 | *SLC25A48* | 5 | 5:135871614:A:G | rs747797136 | 135871614 |
| ENSG00000145832 | *SLC25A48* | 5 | 5:135834851:G:C | - | 135834851 |
| ENSG00000145832 | *SLC25A48* | 5 | 5:135871631:C:G | - | 135871631 |
| ENSG00000145832 | *SLC25A48* | 5 | 5:135871459:A:G | rs200164783 | 135871459 |
| ENSG00000145832 | *SLC25A48* | 5 | 5:135871560:T:A | - | 135871560 |
| ENSG00000145832 | *SLC25A48* | 5 | 5:135874129:A:G | - | 135874129 |
| ENSG00000145832 | *SLC25A48* | 5 | 5:135871710:G:A | rs374679976 | 135871710 |
| ENSG00000145832 | *SLC25A48* | 5 | 5:135852770:G:A | rs372469032 | 135852770 |
| ENSG00000145832 | *SLC25A48* | 5 | 5:135879996:C:T | rs200806048 | 135879996 |
| ENSG00000145832 | *SLC25A48* | 5 | 5:135874141:AG:A | - | 135874141 |
| ENSG00000145832 | *SLC25A48* | 5 | 5:135871709:G:A | rs534483785 | 135871709 |
| ENSG00000145832 | *SLC25A48* | 5 | 5:135834869:G:A | rs369200536 | 135834869 |
| ENSG00000145832 | *SLC25A48* | 5 | 5:135874090:A:T | rs189159333 | 135874090 |
| ENSG00000145832 | *SLC25A48* | 5 | 5:135874084:G:A | - | 135874084 |
| ENSG00000145832 | *SLC25A48* | 5 | 5:135880062:G:A | rs1356589572 | 135880062 |
| ENSG00000145832 | *SLC25A48* | 5 | 5:135880020:C:G | rs767366702 | 135880020 |
| ENSG00000145832 | *SLC25A48* | 5 | 5:135850476:G:A | rs370830017 | 135850476 |
| ENSG00000145832 | *SLC25A48* | 5 | 5:135871497:C:T | rs377170548 | 135871497 |
| ENSG00000145832 | *SLC25A48* | 5 | 5:135852635:G:A | - | 135852635 |
| ENSG00000145832 | *SLC25A48* | 5 | 5:135874039:C:T | rs1156307418 | 135874039 |
| ENSG00000145832 | *SLC25A48* | 5 | 5:135871479:C:T | rs914709129 | 135871479 |
| ENSG00000145832 | *SLC25A48* | 5 | 5:135852809:C:G | rs946309011 | 135852809 |
| ENSG00000145832 | *SLC25A48* | 5 | 5:135852755:G:A | COSV99192265 | 135852755 |
| ENSG00000145832 | *SLC25A48* | 5 | 5:135880023:C:T | - | 135880023 |

**Legend:**

Associations in the table are based on testing of the "HI_mis" mask (see Methods). The respective
*Variant's rank in the set of driver variants determined based on the forward selection approach (
**This is the difference between the P-value when all qualifying variants are aggregated except fo

**GCKD study (HI_mis mask) and their associations with urine choline levels.**

| Non-effect allele | Effect allele | P-value | Effect size | SE | P-value burden test all variants | Rank in set of drivers* |
|---|---|---|---|---|---|---|
| C | A | 4.2E-03 | 2.71 | 0.95 | 1.4E-17 | 5 |
| C | T | 1.2E-03 | 2.16 | 0.67 | 1.4E-17 | 4 |
| A | G | 8.0E-05 | 2.16 | 0.55 | 1.4E-17 | 3 |
| G | C | 2.6E-02 | 2.11 | 0.95 | 1.4E-17 | 6 |
| A | T | 3.6E-02 | 1.99 | 0.95 | 1.4E-17 | 17 |
| G | T | 1.1E-01 | 1.52 | 0.95 | 1.4E-17 | 7 |
| A | G | 1.2E-01 | 1.49 | 0.95 | 1.4E-17 | 8 |
| G | C | 1.3E-01 | 1.44 | 0.95 | 1.4E-17 | 20 |
| C | G | 1.3E-01 | 1.44 | 0.95 | 1.4E-17 | |
| A | G | 3.1E-08 | 1.40 | 0.25 | 1.4E-17 | 1 |
| T | A | 1.8E-01 | 1.27 | 0.95 | 1.4E-17 | 10 |
| A | G | 1.8E-01 | 1.26 | 0.95 | 1.4E-17 | 11 |
| G | A | 2.5E-01 | 1.08 | 0.95 | 1.4E-17 | 12 |
| G | A | 2.9E-01 | 1.00 | 0.95 | 1.4E-17 | 14 |
| C | T | 9.7E-04 | 0.99 | 0.30 | 1.4E-17 | 2 |
| AG | A | 3.0E-01 | 0.99 | 0.95 | 1.4E-17 | 15 |
| G | A | 3.0E-01 | 0.98 | 0.95 | 1.4E-17 | 16 |
| G | A | 1.5E-01 | 0.96 | 0.67 | 1.4E-17 | 9 |
| A | T | 2.1E-01 | 0.69 | 0.55 | 1.4E-17 | 13 |
| G | A | 6.2E-01 | 0.47 | 0.95 | 1.4E-17 | 18 |
| G | A | 6.4E-01 | 0.44 | 0.95 | 1.4E-17 | 19 |
| C | G | 5.1E-01 | 0.44 | 0.67 | 1.4E-17 | |
| G | A | 7.0E-01 | 0.36 | 0.95 | 1.4E-17 | |
| C | T | 9.1E-01 | 0.11 | 0.95 | 1.4E-17 | |
| G | A | 9.1E-01 | 0.11 | 0.95 | 1.4E-17 | |
| C | T | 9.3E-01 | 0.08 | 0.95 | 1.4E-17 | |
| C | T | 7.8E-01 | -0.27 | 0.95 | 1.4E-17 | |
| C | G | 7.5E-01 | -0.30 | 0.95 | 1.4E-17 | |
| G | A | 4.4E-01 | -0.73 | 0.95 | 1.4E-17 | |
| C | T | 3.6E-01 | -0.86 | 0.95 | 1.4E-17 | |

P-value from a burden test of all selected variants using the "LoF_mis" mask with urine ch[illegible]
Bombal et al. 2022 (PMID 35568032)). The greater the variant's contribution to the gene si[illegible]
r the variant in this row itself and the total P-value including all qualifying varaints. The gre[illegible]

| Delta P-value burden test** | Consequence | Protein position | Amino Acids | Codons | cDNA position | CDS position |
|---|---|---|---|---|---|---|
| 1.6E-16 | missense variant | 64 | F/L | ttC/ttA | 377 | 192 |
| 6.3E-16 | stop gained | 243 | R/* | Cga/Tga | 912 | 727 |
| 3.9E-15 | missense variant | 27 | D/G | gAc/gGc | 265 | 80 |
| 7.7E-17 | missense variant | 179 | R/P | cGg/cCg | 721 | 536 |
| 1.0E-17 | missense variant | 159 | Q/L | cAg/cTg | 661 | 476 |
| 3.3E-17 | missense variant | 47 | R/L | cGc/cTc | 325 | 140 |
| 3.0E-17 | missense variant | 192 | Y/C | tAt/tGt | 760 | 575 |
| -1.6E-18 | missense variant | 2 | G/R | Gga/Cga | 189 | 4 |
| -1.6E-18 | missense variant | 198 | P/A | Ccc/Gcc | 777 | 592 |
| 7.9E-12 | splice acceptor variant | - | - | - | - | - |
| 2.1E-17 | missense variant | 174 | L/Q | cTg/cAg | 706 | 521 |
| 2.0E-17 | missense variant | 263 | Q/R | cAg/cGg | 973 | 788 |
| 1.4E-17 | missense variant | 224 | G/D | gGc/gAc | 856 | 671 |
| 1.1E-17 | missense variant | 124 | V/M | Gtg/Atg | 555 | 370 |
| 4.0E-15 | missense variant | 281 | A/V | gCg/gTg | 1027 | 842 |
| 1.1E-17 | frameshift variant | 267 | K/X | aAG/aA | 985-986 | 800-801 |
| 1.1E-17 | missense variant | 224 | G/S | Ggc/Agc | 855 | 670 |
| 2.7E-17 | missense variant | 8 | D/N | Gac/Aac | 207 | 22 |
| 1.3E-17 | missense variant | 250 | Y/F | tAt/tTt | 934 | 749 |
| -3.8E-19 | missense variant | 248 | G/E | gGg/gAg | 928 | 743 |
| -1.0E-18 | missense variant | 303 | R/H | cGc/cAc | 1093 | 908 |
| -2.0E-18 | missense variant | 289 | A/G | gCg/gGg | 1051 | 866 |
| -2.2E-18 | missense variant | 48 | V/M | Gtg/Atg | 327 | 142 |
| -5.3E-18 | missense variant | 153 | A/V | gCg/gTg | 643 | 458 |
| -5.3E-18 | missense variant | 79 | V/I | Gtc/Atc | 420 | 235 |
| -5.5E-18 | missense variant | 233 | T/I | aCa/aTa | 883 | 698 |
| -8.5E-18 | missense variant | 147 | S/F | tCc/tTc | 625 | 440 |
| -8.7E-18 | missense variant | 137 | P/A | Ccg/Gcg | 594 | 409 |
| -1.1E-17 | missense variant | 119 | G/R | Ggg/Agg | 540 | 355 |
| -1.1E-17 | missense variant | 290 | A/V | gCc/gTc | 1054 | 869 |

ɔline levels (analogous to column L) was 4.1e-14, suggesting that the "HI_mis" mask reflected the geneti‹
gnal, the lower the rank-number. Variants without rank are non-drivers. The P-value of the burden test v
ater the variant's contribution to the gene signal, the greater the difference delta.

| Transcript | Impact | REVEL score | CADD PHRED score | M-CAP score |
|---|---|---|---|---|
| ENST00000681962.1 | MODERATE | 0.506 | 23.5 | 0.14684 |
| ENST00000681962.1 | HIGH | | 18.84 | |
| ENST00000681962.1 | MODERATE | 0.808 | 26.3 | 0.437951 |
| ENST00000681962.1 | MODERATE | 0.925 | 22 | 0.179264 |
| ENST00000681962.1 | MODERATE | 0.322 | 16.71 | 0.153748 |
| ENST00000681962.1 | MODERATE | 0.228 | 16.13 | 0.031833 |
| ENST00000681962.1 | MODERATE | 0.79 | 22.4 | 0.156308 |
| ENST00000681962.1 | MODERATE | 0.35 | 24.3 | 0.126892 |
| ENST00000681962.1 | MODERATE | 0.637 | 22.4 | 0.217173 |
| ENST00000681962.1 | HIGH | | 32 | |
| ENST00000681962.1 | MODERATE | 0.883 | 22.3 | 0.167453 |
| ENST00000681962.1 | MODERATE | 0.41 | 19.92 | 0.107926 |
| ENST00000681962.1 | MODERATE | 0.831 | 17.64 | 0.193558 |
| ENST00000681962.1 | MODERATE | 0.537 | 29.4 | 0.0841 |
| ENST00000681962.1 | MODERATE | 0.328 | 18.76 | 0.072994 |
| ENST00000681962.1 | HIGH | | | |
| ENST00000681962.1 | MODERATE | 0.689 | 17.72 | 0.252291 |
| ENST00000681962.1 | MODERATE | 0.424 | 31 | 0.152706 |
| ENST00000681962.1 | MODERATE | 0.335 | 17.61 | 0.066022 |
| ENST00000681962.1 | MODERATE | 0.575 | 18.76 | 0.07625 |
| ENST00000681962.1 | MODERATE | 0.475 | 19.39 | 0.150603 |
| ENST00000681962.1 | MODERATE | 0.655 | 19.26 | 0.060095 |
| ENST00000681962.1 | MODERATE | 0.216 | 14.69 | 0.026396 |
| ENST00000681962.1 | MODERATE | 0.17 | 13.75 | 0.036189 |
| ENST00000681962.1 | MODERATE | 0.358 | 22.6 | 0.042314 |
| ENST00000681962.1 | MODERATE | 0.403 | 18.94 | 0.081387 |
| ENST00000681962.1 | MODERATE | 0.342 | 15.88 | 0.065679 |
| ENST00000681962.1 | MODERATE | 0.179 | 14.63 | 0.055144 |
| ENST00000681962.1 | MODERATE | 0.722 | 22.9 | 0.116162 |
| ENST00000681962.1 | MODERATE | 0.352 | 20.1 | 0.052223 |

c architecture of SLC25A48 better than the variants in the "LoF_mis" mask.
vhen only driver variants are aggregated is 3.4e-21.
